# Phylogeography and transmission dynamics of the 2025–2026 Utah-Arizona measles outbreak: a retrospective genomic epidemiology study

**DOI:** 10.64898/2026.11.10.26367744

**Authors:** Abbey Marye, Kelly Oakeson, Yu Lan, Joshua L. Warren, Drew Francis, Liam Hicks, Katharine S. Walter

## Abstract

**Background:** Measles outbreaks are increasing in frequency globally as vaccine coverage decreases. During the 2025-26 Utah-Arizona border outbreak, identifying where and between whom transmission occurred was essential for directing control measures. However, epidemiological data alone are not sufficient to infer transmission patterns.

**Methods:** As part of the local and state-wide outbreak investigation, we conducted amplicon-based whole genome sequencing on residual diagnostic samples from confirmed measles cases collected from June 15, 2025–April 30, 2026. We reconstructed viral dispersal across Utah using Bayesian phylogeographic analyses. We evaluated genetic distance thresholds for identifying recent transmission by comparing single nucleotide polymorphism (SNP)-based genetic clusters with known epidemiologic linkages. We then assessed pairwise exposures associated with genetically supported transmission linkage using logistic regression, accounting for network and spatial dependence between pairs.

**Findings:** We generated 509 high-quality whole measles virus genomes from 913 patients. In 2025, most viral dispersal events (67.9%) originated in Southwest Utah and Mohave County, Arizona, where the first cases were reported. By 2026, most viral spread (60.5%) occurred outside of this area. A threshold of 0 SNPs identified epidemiologically linked pairs of individuals with a sensitivity of 77.4% and specificity of 92.3%. Household membership (odds ratio (OR): 219.10, 95% credible interval (CI): 112.66-325.53), participation in the same sport (OR: 15.19, 95% CI: 9.75-20.63), and attendance at the same school (OR: 51.5, 95% CI: 36.38-66.61) were independently associated with transmission linkage. Increasing distance between households was associated with reduced odds of linkage (OR: 0.46: 95% CI 0.44-0.48).

**Interpretation:** Repeated introductions of measles virus from southwestern Utah seeded transmission elsewhere in Utah, with household, school, and sports exposures contributing to onward transmission. Validated genome-based linkage measures can complement contact tracing by helping public health teams identify likely transmission networks and prioritize investigation when resources are constrained.

**Funding:** Epidemiology and Laboratory Capacity (ELC) program at the CDC

**Research in Context:** *Evidence before this study:* We searched PubMed and medRxiv as of October 6, 2026 for studies about measles genomic or molecular epidemiology, measles virus transmission modelling, and application of whole genome sequencing of measles virus. English language search terms included “measles”, “measles virus”, “phylodynamic”, “genomic epidemiology”, “molecular epidemiology”, “transmission modelling”, “transmission dynamics”, “sequencing”, “whole genome sequencing”, and “WGS”. Previous studies have primarily utilized N450 sequencing in their outbreak assessments, including phylogenetic and phylogeographic analyses. More recent studies have utilized whole genome sequencing to perform phylogenetic analysis and have overlaid some epidemiological data to assess transmission. To our knowledge, no studies have combined epidemiological and whole genome sequencing data to quantitatively assess drivers of transmission.

*Added value of this study:* We integrate detailed epidemiological data with whole genome sequencing, providing a comprehensive analysis of geographic spread and key drivers of transmission in the Utah-Arizona border outbreak. We generated whole genome sequences for 56% of confirmed cases, high sequence coverage that enabled us to identify an appropriate genomic threshold and approximate direct transmission links. Our findings illustrate a change in geographic transmission as the outbreak persisted and highlighted household, school, and sports based transmission as primary sources of exposure.

*Implications of all the available evidence:* Our findings demonstrate the utility of measles surveillance using whole genome sequencing paired with epidemiological data. As measles outbreaks increase in frequency, new approaches are needed to characterize the key drivers of transmission. By integrating epidemiologic and genomic data, we identified genomic thresholds for transmission that can be applied in settings where extensive epidemiologic data are unavailable. Ultimately, utilizing genomic and epidemiological data together will strengthen our surveillance of future measles outbreaks.

## Introduction

Measles resurgence threatens its elimination in the United States and globally.^1–3^ Measles is among the most transmissible infectious diseases; in populations with gaps in vaccination, a single introduction can lead to rapid and prolonged transmission. As outbreaks grow, conventional epidemiologic case investigation becomes difficult. Identifying where and between whom measles virus transmission occurs, and how the virus is introduced into new locations is essential for blocking transmission and ending outbreaks. Traditional analysis of measles outbreaks often rely on mathematical models and epidemiological data to determine transmission drivers or deduce spatio-temporal transmission patterns.^4,5^ However, genomic data offer an added layer of resolution to better understand transmission.

Routine molecular surveillance for measles relies on sequencing a 450-nucleotide region of the nucleoprotein gene (N-450), which is used to assign genotypes.^6^ Molecular epidemiology can complement conventional epidemiological investigation, but thus far, its resolution for measles virus has been limited.^7,8^ However, declining global measles virus diversity and the predominance of only a few circulating genotypes means that epidemiologically unrelated cases can share identical N-450 sequences.^9^ Whole genome sequencing provides greater resolution and has been used to identify viral introductions, epidemiologically defined clusters, and reconstruct transmission chains.^10,11^

Previous measles genomic studies have principally used epidemiologically defined groups to interpret phylogenetic structure or to reconstruct plausible transmission linkages. For example, whole-genome sequences have clarified transmission following the 2010 Winter Olympic Games in Vancouver, distinguished epidemiological clusters in the Netherlands, and separated transmission chains among Afghan evacuees housed in congregate settings in the United States.^12–14^ However, to our knowledge, no previous study has empirically validated a genomic approach for inferring recent measles transmission against a large database of known epidemiological links, nor used genome-supported linkage to quantify the relative importance of specific exposures to transmission.

In August 2025, measles was reported in insular communities spanning Mohave County, Arizona, and southwest Utah.^15^ Members of this community cross the state border frequently, often attending school or working on one side and living on the other. Transmission subsequently expanded across Utah and, as of September 2026, it is the longest sustained individual outbreak in the United States in the twenty-first century, involving more than 1000 reported cases.^3,16–19^ The geographic expansion and prolonged transmission highlights the urgency of understanding how the virus moved from the border region across the state, and of determining the types of contacts responsible for transmission.

We integrated whole-genome sequencing with detailed epidemiologic and contact tracing data from confirmed Utah cases. We aimed to reconstruct the spatial dispersal of measles virus across the state; evaluate genomic-distance criteria for identifying epidemiologically linked cases; and quantify associations between household, school, sporting, workplace, and geographic exposures and genome-supported recent transmission. By densely sampling a large, prolonged, community outbreak, we sought to establish a generalisable framework for using measles virus genomic epidemiology to complement contact tracing when observed transmission networks are incomplete.

## Methods

### Whole Genome Sequence Data and Processing

Measles virus PCR–positive clinical specimens submitted to or tested at the Utah Public Health Laboratory (UPHL) were whole genome sequenced using the Artic Measles 400 bp v1.0.0 primer set from Primal Scheme on the Illumina NextSeq platform.^20^ Raw reads were processed and consensus sequences were generated using the Cecret pipeline.^21^ The Cecret pipeline includes quality filtering (seqyclean), alignment (bwa), primer trimming (ivar), and consensus genome generation (ivar).^22–24^ We performed taxonomic classification using Kraken2 on the reads. Raw sequence data have been deposited in the NCBI Sequence Read Archive under the BioProject accession number PRJNA1293457.

### Case and Sequence Inclusion

Cases met the criteria for inclusion if they had an onset date or sample collection date during June 15, 2025–April 30, 2026. This start date was chosen because cases with onset dates at this time were confirmed to be genomically related and linked to the outbreak. The selected end date allowed for all case investigations to be completed and sequencing to be completed before beginning data analysis. Any case from Utah or Mohave County, Arizona was included. Since the majority (97%) of cases tied to the outbreak from Arizona resided in Mohave County, all cases from other jurisdictions in Arizona were excluded. Cases that were not in the same monophyletic clade as cases from the outbreak or its continued transmission were also excluded (21 cases). Furthermore, sequences were required to have ≥90% consensus coverage and ≤25% ambiguous bases to be included for analysis.

### Bayesian Phylogenetic Analysis and Ancestral State Reconstruction

Each case was assigned a date corresponding to either rash onset date, or sample collection date if rash onset date was not available. Each case was assigned a local health district (location) based on the address at diagnosis in the Utah disease surveillance database. ClustalW sequence alignments were created using Clustal Omega and were then imported into Beauti, along with dates and location information.^25^ We specified an HKY substitution model, a strict molecular clock rate of 0.000661 (substitutions per site/year), and a Coalescent Constant Population prior.^26,27^ We fit a Bayesian timed tree in BEAST2 (v2.7.7), running a Markov chain Monte Carlo posterior sampling algorithm for 10,000,000 iterations.^25^ We used a Markov Jump analysis to calculate the number of jumps between locations and used R to complete further analysis of viral introductions.^28^

### Genomic Cluster Analysis

SNP distance matrices were generated using the snp-dists package in Python.^29^ We then applied SNP-based genetic distance thresholds to cluster isolates using the package transcluster, with the function makeSNPClusters in R.^30^ This package uses a SNP distance matrix to infer genomic clusters. A SNP threshold is then chosen as a cutoff for identifying genomes likely related by recent transmission. To assess concordance between the epidemiological and genomic clustering, we calculated the Adjusted Rand Index at SNP thresholds of 0–6 SNPs).^31^ We assessed the sensitivity and specificity of our genomic clustering approach, across the different SNP thresholds (0–6), using observed epidemiological links as a gold standard. Epidemiological links are defined as cases with verified contact with another measles case within the viral incubation period.^32^

### Transmission Driver Analysis

We quantified associations between epidemiological exposures and pairwise genomic clustering, as a proxy for recent transmission, using GenePair in R.^33^ GenePair is a hierarchical Bayesian logistic regression model that accounts for network and spatial correlation between spatially-referenced pairs in a transmission chain which allows for more accurate statistical inference when estimating the effect of factors on genomic clustering.^34^ We chose six pairwise exposure variables (shared household, shared school, shared sport participation, shared age group, shared zip code, and shared local health district) to assess the extent to which they drive transmission both within and between local health districts. Shared household was defined as individuals sharing the same primary physical living space or physical address; shared school was defined as individuals reporting attendance at the same school; shared sport participation was defined as individuals who report participating in the same sport (not necessarily the same team); and shared age group was defined as individuals within the same age bracket (<5 years old, 5–13 years old, 14–19 years old, 20–39 years old, 40–59 years old, >60 years old). The age brackets were chosen to capture children <19 years old who were in contact in other ways outside of school, and to capture potential contacts in adult work environments. We verified that there was no unexpected collinearity between our chosen variables (Table SI 1).

Missing data in shared household, shared school, and shared sport participation were accounted for. Pairs of cases where one member was under 18 years old, and one was over were assumed to not share a school exposure. While there are adults present in school settings, epidemiological investigation often extracted employment information, so we assumed that all adult school exposures were accounted for. Additionally, since only ∼1% of students commute more than 30 miles to attend school, we assumed that any pair of cases more than 60 miles apart were not likely to have the same school exposure.^35,36^ Using these assumptions, we were able to verify the shared school status of 88.8% of the pairs in our data. Since sport participation is state-wide, we only used the age difference assumption to verify the shared sport status. We also assumed that only children within the same age group would be participating in sports together. Using this assumption, we were able to verify the shared sport status of 83.9% of the pairs in our data. There were 17 individuals whose age was not obtained in the investigation. After filtering for these individuals, we were able to verify the shared age group of 93.4% of the pairs in our data. If shared household information was not known for a pair, the distance between cases was used to determine shared household status. We took the median distance between pairs that shared a household, which was 28 miles. If cases were more than 28 miles away from each other, they were considered to not share a household. Using this assumption, we were able to verify 93.1% of the pairs in our data. Only the pairs where shared household, shared school, shared sport, and shared age group could be verified (76.1% of original pairs) were included in the analysis. There was no missing data for zip code or local health district (LHD).

Our analysis utilized 98,407 pairs from 492 samples after accounting for missing data. We specified independent prior distributions for the regression parameters as □*_j_* ∼ N(0,1.00) while all other parameters remained as the model’s defaults (i.e., weakly informative inverse gamma priors for variance parameters and a gamma prior distribution for the spatial correlation parameter). We collected 90,000 posterior samples in total, removing the first 1,000 iterations as burn-in from each of ten independent parallel chains that were each run for 10,000 iterations. The average effective sample size across each variable was > 7000 and convergence was assessed by visual inspection of traceplots for all key model parameters and calculating the Gelman-Rubin statistics for each of the regression parameters (average R = 1.00).^37^

## Results

### Geographic Transmission of Measles Virus in Utah and Arizona

By April 30, 2026 there had been 913 cases of measles associated with the outbreak that originated on the Utah-Arizona border. While the majority of cases originated from the two LHDs that straddle the border (Southwest Utah and Mohave County), by the end of February 2026 there had been confirmed cases in every LHD in Utah. We were able to obtain whole genome sequencing for 509 (56%) of these cases, originating from every LHD except San Juan.

To reconstruct measles virus spread across Utah, we conducted ancestral state reconstruction to infer the likely geographic location of ancestral viral lineages. In 2025, we found that viral movement was largely (71%) between the two health districts straddling the Utah-Arizona border (Southwest Utah and Mohave County). In 2026, most viral movement (74%) was between LHDs in Utah other than Southwest Utah. However, since 87% of the introductions into either Southwest Utah and Mohave County can be attributed to transmission between those two LHDs, we combined them into one LHD for the remainder of the analyses (Figure 1).

**Figure 1:**
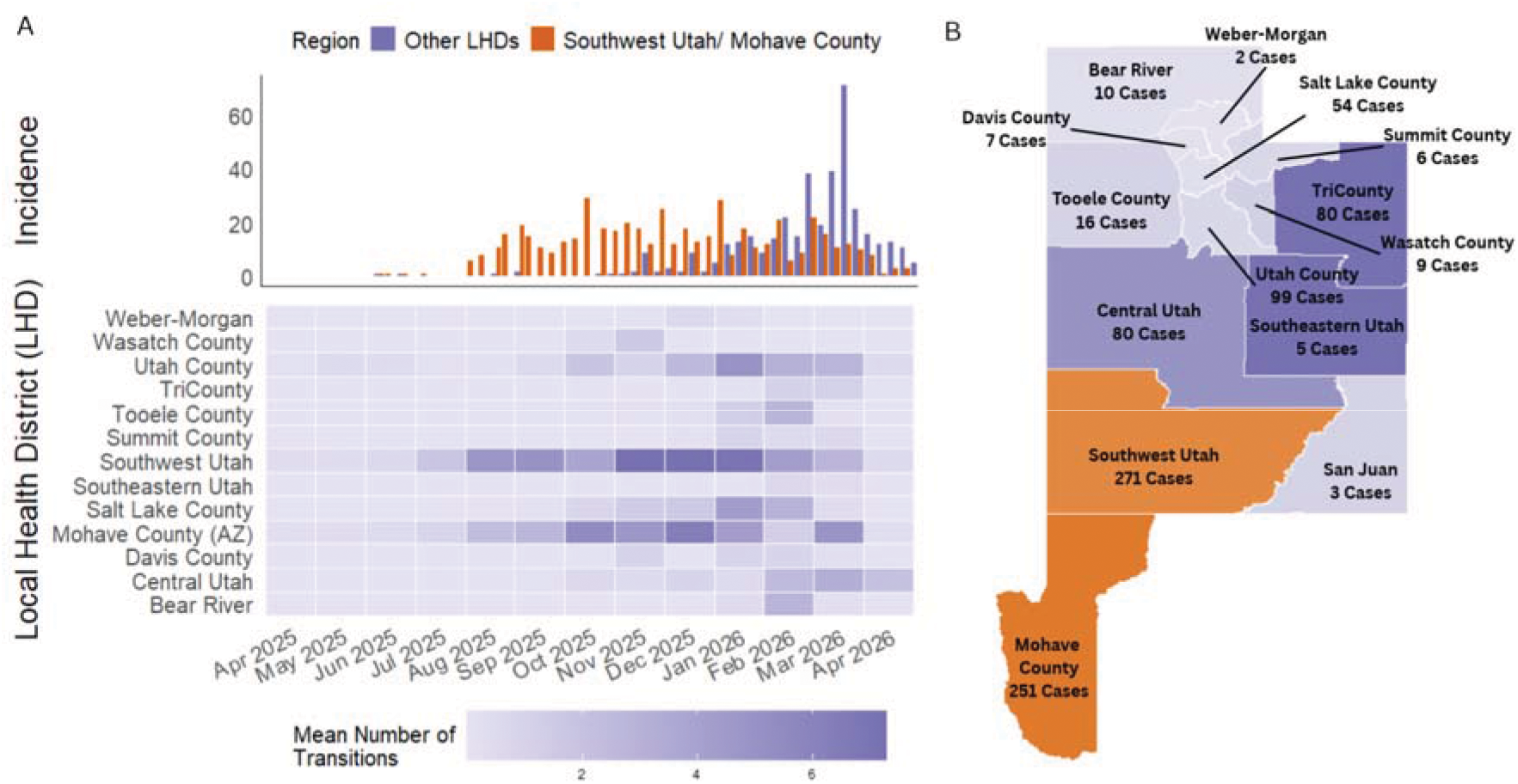
Introductions of measles virus across Utah. (A) Incidence curve showing the number of confirmed weekly measles cases from June 13, 2025 - April 30, 2026 colored by whether the case occurred in Southwest Utah/Mohave County or another local health district (LHD) in Utah. The number of transitions (mean Markov jumps) of measles virus into each LHD from any other LHD are shown on the same temporal axis below. (B) The total number of cases in each LHD as of April 30, 2026 colored by either being Southwest Utah/Mohave County or being another LHD in Utah.

In 2025, many cases (34%) seen outside of Southwest Utah/Mohave County reported an epidemiological link or travel history to the region, and ancestral state reconstruction estimated that, in 2025, 62.7% of introductions to the seven LHDs with confirmed cases occurred from Southwest Utah/Mohave County. In contrast, in 2026, every LHD in Utah had at least one confirmed case, many of which had no reported epidemiologic link to Southwest Utah/Mohave County. Ancestral reconstruction is consistent with this, showing only 39.1% of estimated transmissions originating from Southwest Utah/Mohave County to the ten LHDs that had sequenced confirmed cases (Figure 2).

**Figure 2:**
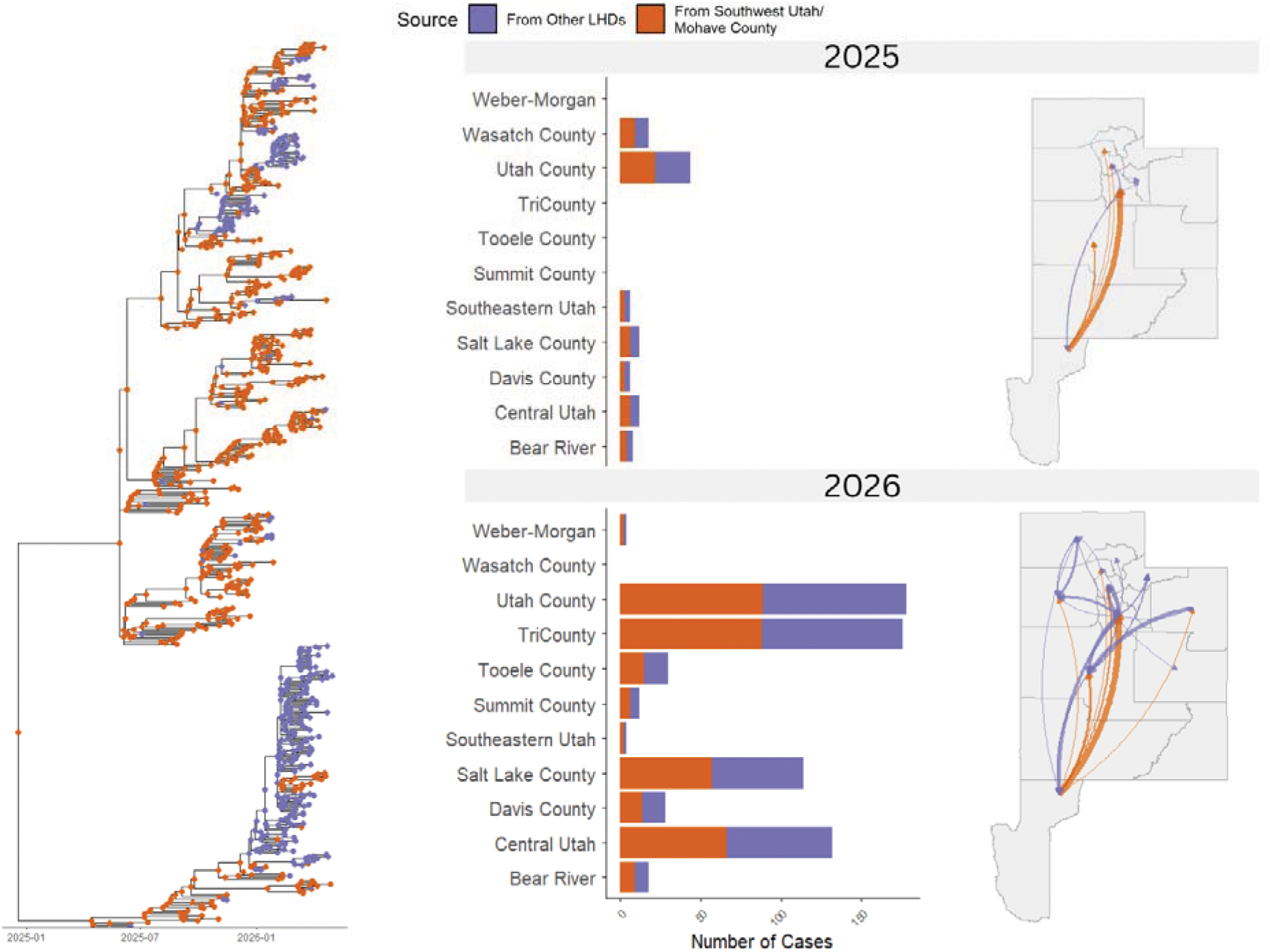
Geographic spread of measles virus across Utah. On the left, a time-scaled phylogenetic tree is shown with the tips/nodes colored by actual or inferred geographic location in either Southwest Utah/Mohave County or any other LHD. In the center, the number of transitions (mean Markov jumps) from Southwest Utah/Mohave County vs. from other LHDs to a given LHD is shown in stacked barplots. On the right, arrow weight indicates the number of inferred introductions between LHDs, and arrow color indicates viral source, with panels indicating year.

To further investigate this change, we determined which LHDs were the primary sources of transmission to other LHDs besides Southwest Utah/Mohave County. We found that in 2025, Utah County was the source of 26.1% of introductions to other LHDs and 40.5% of introductions into other LHDs in 2026. In 2026, Utah County accounted for the most transmissions (40.5%) to other LHDs, more than Southwest Utah/Mohave county (39.1%).

We predicted that after the virus was introduced across the state from Southwest Utah/Mohave County, local transmission sustained spread. In this situation, we would expect that pairwise genetic distances would be correlated with pairwise geographic distance between patient zip codes. Our analysis shows that there was moderate correlation (r^2^ = 0.33) between the SNP distance and geographic distance between cases. The correlation was weaker when pairs were analyzed separately between LHDs (r^2^ = 0.27) and within LHDs (r^2^ = 0.19). We also found that there was little change in this correlation over time when we compared 2025 cases (r^2^ = 0.15 (between), r^2^ = 0.24 (within)) with 2026 cases (r^2^ = 0.20 (between), r^2^ = 0.20 (within)) (Figure 4). Overall, genetic distance was more closely correlated with geographic distance within LHDs as compared to between LHDs, consistent with a pattern of geographic distance partly explaining local spread, and being less important in long-distance viral movement.

### Estimation of Genomic Clustering Using a SNP Threshold

For many pathogens, pairwise genomic distances are frequently used as a proxy for recent transmission and therefore, to identify transmission linkages. We evaluated alternative pairwise distance thresholds to determine whether SNP-based clustering could provide a consistent proxy for epidemiological linkage, as determined by contact tracing. Applying a pairwise genetic distance threshold of 0 SNPs yielded 146 genomic clusters and recovered known epidemiologically linked pairs with a sensitivity of 77.4% and specificity of 92.3% (Figure 3).

**Figure 3:**
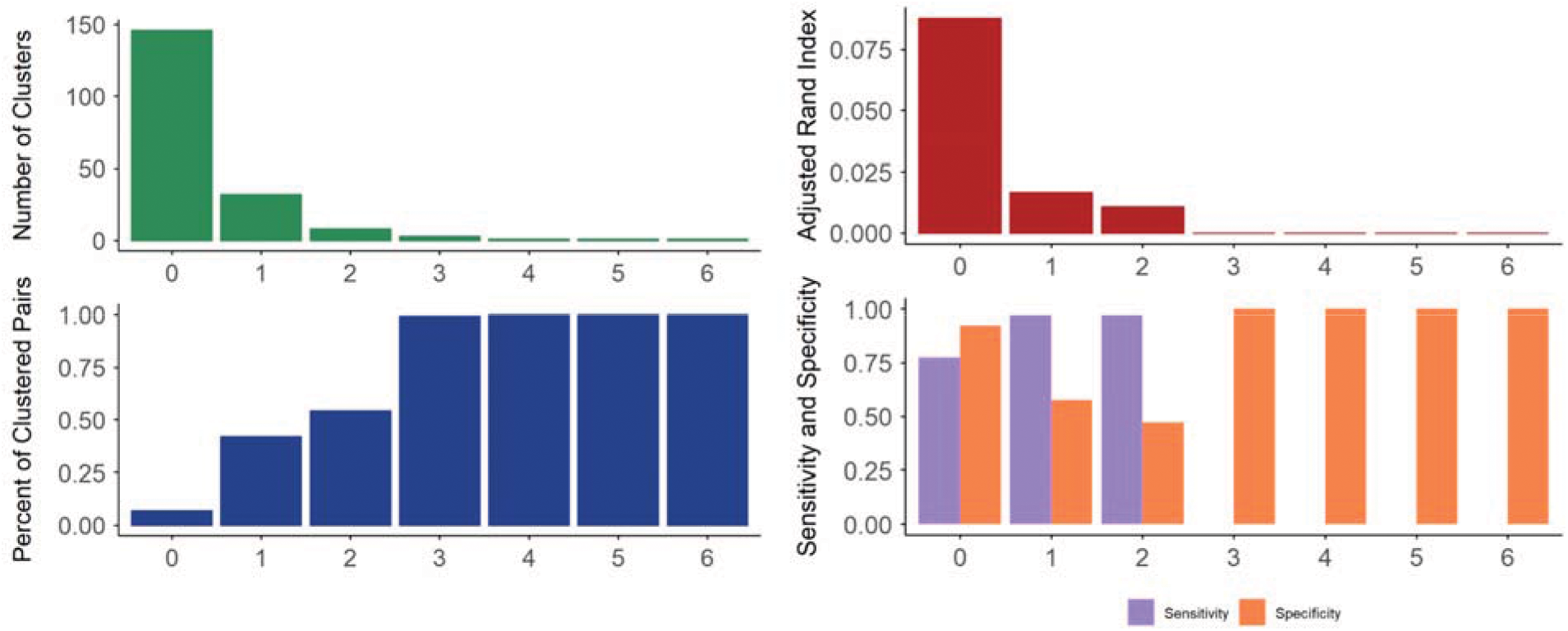
Performance of genetic distance thresholds in recovering known epidemiologic linkages: (A) The number of genomic clusters identified. (B) The Adjusted Rand Index between the genomic clusters created and the epidemiological clusters assigned through contact tracing. (C) The percent of pairs that fall within a genomic cluster. (D) The sensitivity and specificity of the genomic clusters created versus the epidemiological clusters.

### Analysis of Primary Transmission Drivers

The outbreak investigation conducted at the Utah DHHS consisted primarily of case interviews. During these interviews, epidemiologists gathered epidemiological and demographic information related to possible exposures. To identify the types of exposures driving transmission, we tested whether specific pairwise exposures (shared household membership, shared school affiliation, shared sport affiliation, shared LHD, shared age group, and geographic distance) were associated with genomic clustering (Table 1) using a 0 SNP threshold, as a proxy for transmission.

**Table 1:**
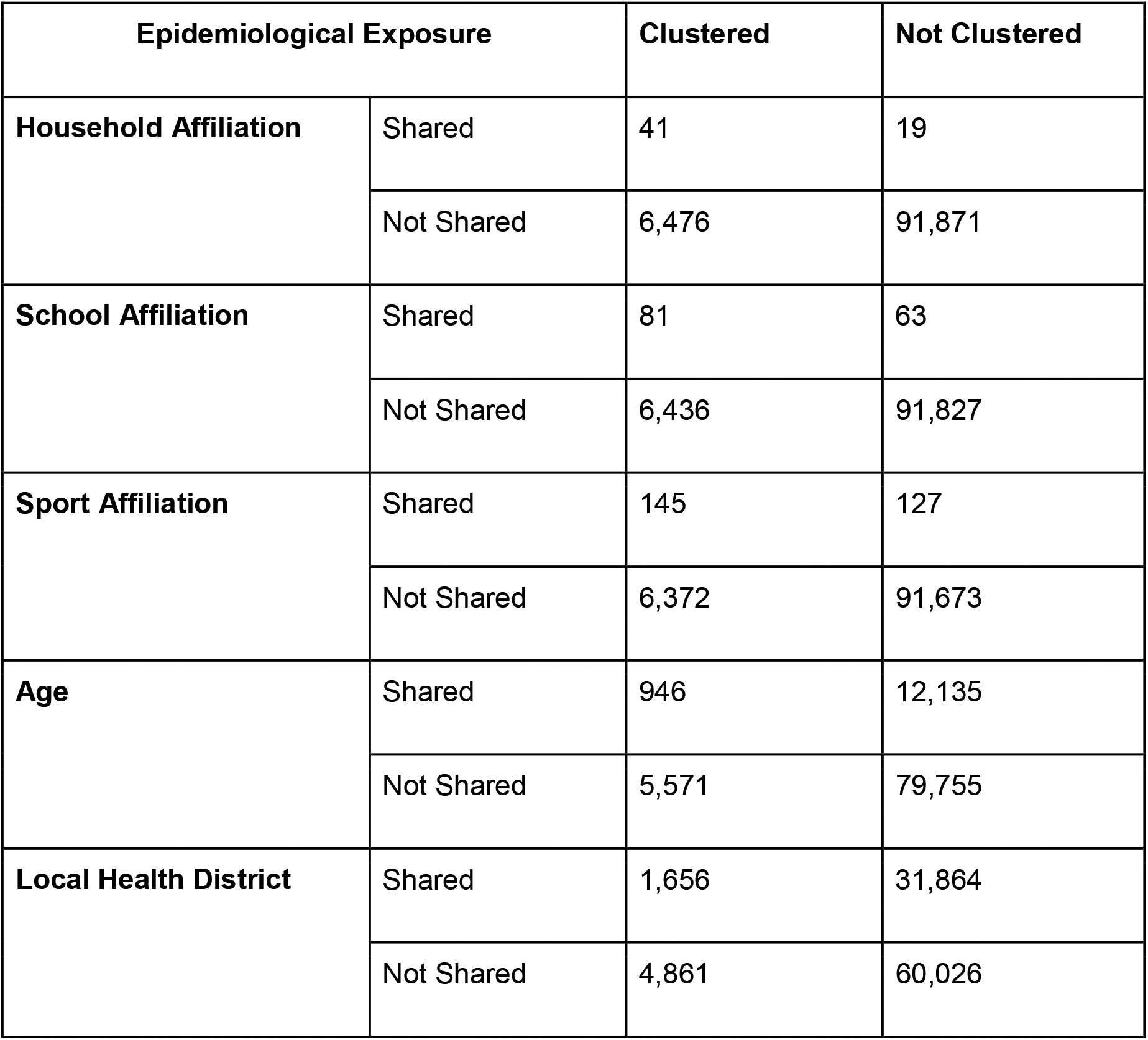
Pairwise epidemiological exposures and genetic clustering. Two-way tables of variables used in transmission driver analysis. The number of pairs with a shared epidemiological exposure vs. without a shared epidemiological exposure are shown by whether they are genomically clustered or not.

In a logistic regression model, which controlled for non-independence of pairs, shared household membership, shared school affiliation, and shared sport participation were all associated with genomic clustering. Cases who shared a household had an increased odds of being genomically clustered (odds ratio (OR) = 219.10, 95% credible interval (CI) 112.66-325.53), cases who shared a school had an increased odds of being genomically clustered (OR = 51.5, 95% CI: 36.38-66.61), and cases who participated in the same sport had an increased odds of being genomically clustered (OR = 15.19, 95% CI: 9.75-20.63) (Figure 4). While distance between households was associated with a reduced odds of genomic clustering (OR = 0.46, 95% CI: 0.44-0.48), cases who live in the same LHD have an increased odds of being genomically clustered (OR = 1.99, 95% CI: 1.75-2.24) (Figure 4).

**Figure 4:**
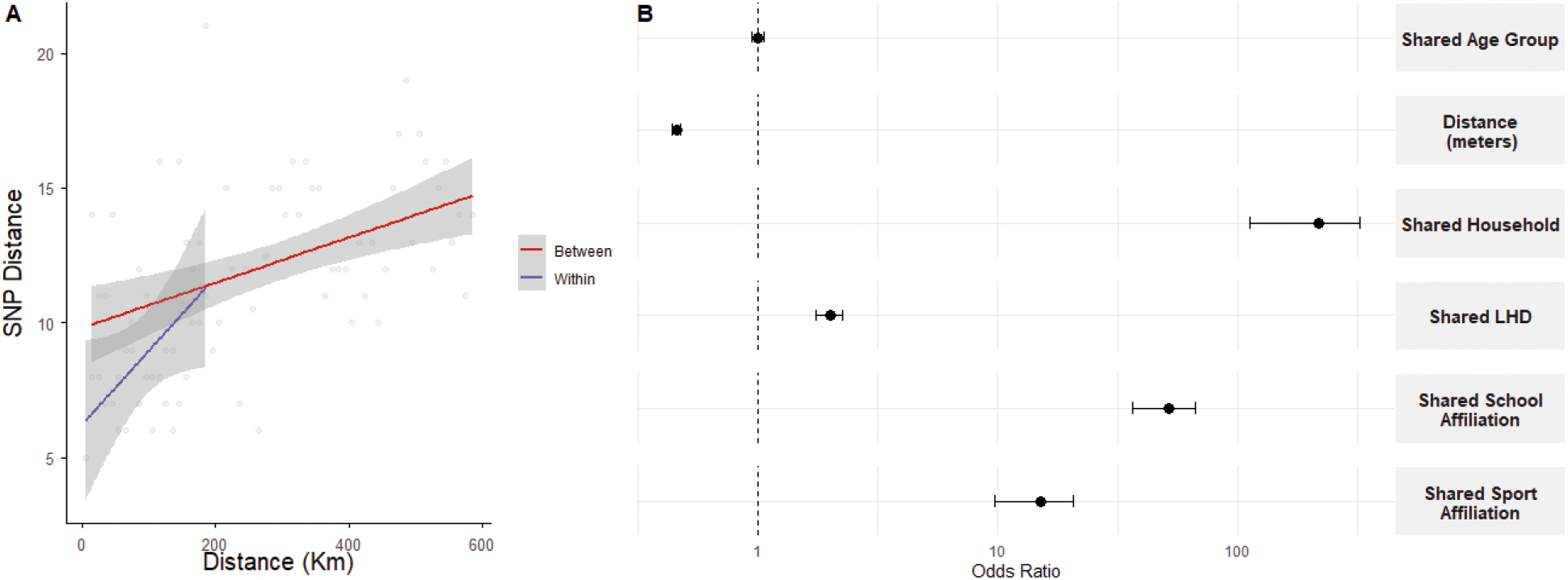
Shared exposures associated with measles virus genomic clustering, as a proxy for recent transmission. (A) Correlation between pairwise measles virus SNP distance and geographic distance in meters plotted for pairs within and between LHDs. Points indicate pairwise correlations, and blue and red lines indicate correlation within the same LHD and in different LHDs, respectively. (B) Odds ratios estimated for the association of pairwise exposures (shared household, school, sport, age group, LHD, and geographic distance) and genomic clustering. Odds ratios are plotted on a logarithmic scale with 95% credible intervals.

## Discussion

By integrating high-coverage measles virus whole-genome sequencing with detailed surveillance data, we characterised how the geographic structure and epidemiological context of measles transmission changed during a prolonged measles virus outbreak in Utah. Early viral dispersal was concentrated around the initially affected Utah–Arizona border region, whereas later transmission was increasingly distributed across Utah. We found that a 0-SNP threshold identified epidemiologically linked case pairs with high specificity but incomplete sensitivity, illustrating both the value and limitations of genomic similarity for identifying recent transmission. Household membership, school attendance, and participation in the same sport were strongly associated with genome-supported linkage. Together, these findings show how measles virus genomic data can complement incomplete contact investigations and identify settings to prioritize for outbreak control.

Traditional analysis of measles outbreaks has mainly relied on N450 sequencing to assess transmission and some previous studies have additionally utilized whole-genome sequencing (WGS) for this purpose. For example, a study assessing transmission in Israel during 2018–2019 was able to obtain WGS for 3% of confirmed measles cases and used that information to determine that their transmission resulted from multiple parallel introductions.^38^ Similarly, a study in the Netherlands was also able to obtain WGS for 3% of their confirmed cases to demonstrate that there was significant genomic clustering without epidemiological links.^13^ Our study builds upon this prior work. We obtained WGS for a large portion (56%) of confirmed cases in a long outbreak, and integrated genomic data with detailed epidemiologic data, enabling greater insight into the changing transmission patterns throughout the duration of the outbreak. We conclude that transmission patterns in the state of Utah changed as the outbreak progressed. Our phylogenetic analysis inferred many probable instances where there was transmission between LHDs. We also found that transmission, or “seeding,” from Southwest Utah/Mohave County to other LHDs occurred more frequently in the first six months of the outbreak than the following four months. This is in contrast to two other large measles outbreaks that occurred during 2025–2026 in the United States in Texas and South Carolina where the majority cases occurred in the health district where the outbreak originated.^39,40^ These findings would not be possible from just traditional epidemiologic data alone, and show that genomic data can allow for more comprehensive understanding of outbreak transmission dynamics.

In addition to delineating how transmission patterns changed over time, we were also able to determine the factors driving transmission. This level of detail has historically not always been available in measles investigations, but has been possible in investigations of pathogens like tuberculosis.^14,41^ We hypothesized that cases that are in close geographic proximity are likely to be genetically related. Our analysis of genomic clusters was consistent with this hypothesis: pairs that were geographically closer had a higher odds of being in the same genomic cluster. We also found that specific types of contacts greatly increased the odds of genomic clustering. Sharing a household, school, or sport was associated with a higher odds of being within the same genomic cluster.

This study has some limitations. While we were able to obtain whole genomes from the majority of our confirmed cases, only cases who were diagnosed via a polymerase chain reaction test were eligible for whole-genome sequencing so we were not able to obtain all of them. This means that the sequenced samples may not be representative of the entire outbreak. Additionally, we were not able to obtain the household affiliation, school affiliation, and sport affiliation for all sequenced cases. Our ability to obtain this epidemiological data was dependent on how much information an interviewed case was willing to give. This willingness is likely not uniform across time or geographic area. We attempted to account for this missingness by using the informed assumptions mentioned in our methods to determine which pairs likely did not share the same affiliations, but we cannot impute shared affiliations. Additionally, this is an analysis from a single outbreak and the genomic thresholds we determined must be validated elsewhere. These limitations highlight the need for increased whole genome sequencing efforts during large outbreaks and for smoother systematic integration of epidemiological and genomic data.

In conclusion, our findings underscore the utility of WGS paired with epidemiological data when investigating a measles outbreak. Dense measles virus sequencing, interpreted alongside exposure and surveillance data, revealed changes in the geographic structure of the Utah-Arizona outbreak and identified specific exposures associated with recent transmission. Although the genomic-linkage criterion requires validation in other outbreaks and viral lineages, our findings provide a framework for integrating WGS with routine measles surveillance in a highly vaccinated population when contact networks are incomplete. Furthermore, with better defined genomic clusters we exemplify how logistic regression accounting for network and spatial dependence within these clusters can be used to analyze drivers of transmission. As measles virus outbreaks increase in frequency, these methods can further our understanding of measles transmission and enhance public health action.

## Data Availability

Raw sequence data have been deposited in the NCBI Sequence Read Archive under the BioProject accession number PRJNA1293457. All consensus sequences are available on NCBI GenBank or Pathoplexus.

https://www.ncbi.nlm.nih.gov/bioproject/PRJNA1293457

## Contributors

AM and KW conceptualized the study design, developed the methods, and created an analysis plan. AM performed the data analysis with assistance from JLW, YL, and KW. AM and KW wrote the first draft of the manuscript with support from KO, DF, and LH. All authors contributed to, reviewed, and approved the final version of the manuscript.

## Ethical Approval

This study was conducted in accordance with the Declaration of Helsinki. As this study used pathogen genomic data obtained through routine public health surveillance and anonymized patient demographic data, formal ethical approval was not necessary.

## Acknowledgments

This study was funded by the Epidemiology and Laboratory Capacity (ELC) program at the CDC Cooperative Agreement Number CK24-0002. All data was generated from the Utah Department of Health and Human Services and was analyzed in collaboration with the Department of Epidemiology at the University of Utah. This work would not be possible without the work of epidemiologists at each local health department in Utah and the Mohave County Department of Public Health.

