## Supplemental Table 1 for "Phylogeography and transmission dynamics of the 2025–2026 Utah-Arizona measles outbreak: a retrospective genomic epidemiology study"

|  | **Household Affiliation** | **School Affiliation** | **Sport Affiliation** | **Age** | **Distance** | **Local Health District** |
| --- | --- | --- | --- | --- | --- | --- |
| **Household Affiliation** | - | - | - | - | - | - |
| **School Affiliation** | 0.09597951 | - | - | - | - | - |
| **Sport Affiliation** | -0.001300372 | 0.01823873 | - | - | - | - |
| **Age** | -0.001183051 | 0.02416365 | 0.04152986 | - | - | - |
| **Distance** | -0.03519779 | -0.05376705 | -0.009792911 | -0.03344921 | - | - |
| **Local Health District** | 0.03262816 | 0.05326146 | -0.02967128 | 0.03393327 | -0.7893564 | - |

**Table SI 1: Correlation coefficients between each variable included in linear regression.**
